# Genetic and Biological Pathways Underlying the Relations between Executive Functioning and Externalizing Behaviors

**DOI:** 10.64898/2026.10.08.26365006

**Authors:** Kellyn M. Spychala, Alex P. Miller, Naomi P. Friedman, Ian R. Gizer

## Abstract

**Background:** Across phenotypic and genotypic studies, executive functioning (EF) has been shown to be associated with externalizing behaviors, including attention-deficit/hyperactivity disorder (ADHD), aggression, and substance use, and is a proposed underlying mechanism of these behaviors. However, there is limited research on the shared biological mechanisms underpinning EF and externalizing behaviors, including their overlap at the level of genomic regions.

**Methods:** The current study used genome-wide association study summary statistics of EF, ADHD, aggressive behavior, alcohol use, and alcohol use disorder to examine whether the heritability of these traits or the co-heritability between them is enriched for genes expressed in particular tissues or stratified by other annotation types. Additionally, the study examined the genomic regions contributing to the covariation between EF and the externalizing traits to identify genetic variants and genes that may be underlying their overlap.

**Results:** First, across all phenotypes, there was evidence for enrichment of genetic signals in DNase hypersensitive sites and H3K4me1 regions, and significant enrichment among genes expressed in the anterior cingulate cortex and frontal cortex. Second, when combining results across regional methods, there was evidence for associations between EF and each of the externalizing behaviors in regions on chromosomes 1, 11, and 15.

**Conclusions:** These findings highlight biological pathways that may <u>link</u> the relations between EF and externalizing behaviors and provides additional evidence of EF as an underlying mechanism of externalizing behavior.

## Background

Childhood and adolescent behaviors, traits, and psychological disorders involving difficulties with self-regulation and rule-breaking, such as aggression, substance use, and attention-deficit/hyperactivity disorder (ADHD), are prevalent (e.g., 3% for ADHD [Polanczyk et al., 2015] and 26% for alcohol use [Volkow et al., 2021]) and *co-occur* (Krueger et al., 2007). Latent variable modeling has demonstrated that the covariance among these phenotypes, characterized by behaviors that violate societal norms (Kauten & Barry, 2020), can be explained by a single factor, which has been theorized to reflect an externalizing spectrum of behavior (Achenbach & Edelbrock, 1991; Krueger et al., 2005). Based on such evidence, it is assumed that these behaviors share an underlying etiology.

Deficits in executive functioning (EF) are hypothesized to represent one shared mechanism involved in the etiology of externalizing behavior (Beauchaine et al., 2014). Deficits in EF have been associated with increased risk for ADHD (Townes et al., 2023; Yang et al., 2022), conduct disorder (Figueiredo et al., 2022), and greater substance use (Castellanos-Ryan & Conrod, 2020; Gustavson et al., 2017). Additionally, longitudinal studies have suggested that stability in externalizing behavior across development can be partially accounted for by EF deficits (Papachristou & Flouri, 2020). Equally important, however, are studies suggesting that an externalizing factor explains only a portion of the overall variance in the constituent behaviors and other research suggesting there may be unique relations between EF and individual externalizing behaviors (Vassileva & Conrod, 2019). For example, Castellanos-Ryan et al. (2014) reported distinct patterns of EF deficits and associated fMRI response for ADHD and conduct disorder relative to substance use disorders (SUDs) where ADHD and conduct disorder were associated with errors on a continuous performance task, but substance misuse was not. Such results illustrate the importance of examining the relations between EF and externalizing behavior at both the shared and behavior-specific levels.

EF processes have been localized, in part, to the prefrontal cortex (PFC) and the anterior cingulate cortex (ACC) regions of the frontal lobe. Localized activation of the dorsolateral (DL) PFC and connectivity between this area and the ACC increases as EF develops across the lifespan, especially during early and middle childhood (Fiske & Holmboe, 2019). Further, when EF tasks include a motivational component, neuroimaging studies have shown engagement of the DLPFC as well as the medial and orbital PFC and the ventral ACC (Salehinejad et al., 2021). Among individuals exhibiting externalizing behaviors, decreased activation of the ACC and PFC, including the medial PFC, compared to healthy controls has been demonstrated (Heinz et al., 2011; Sterzer et al., 2005; Yasumura et al., 2019). Additionally, animal studies have shown that dopaminergic systems regulate glutamatergic transmission within PFC neurons (Yuen et al., 2010), and alterations in glutamate transmission can impair aspects of EF, specifically working memory (Aultman & Moghaddam, 2001). These results overlap with prior studies demonstrating relations between dopaminergic and serotonergic neurotransmission in the PFC with ADHD and aggression, respectively (Yan & Rein, 2022 and Davidson et al., 2000).

The reviewed findings highlight *overlap* in the neurobiology of EF and externalizing behaviors, but more recent studies have begun to test the shared *directional* relations between these neurobiological underpinnings and both EF deficits and externalizing behaviors (Poletti, 2009). For example, EF deficits and SUDs have been shown to covary with structural differences in the brain, including reductions in grey matter volume of the ACC (Goodkind et al., 2015). Additionally, impairments in EF can be exacerbated by disruptions of dopaminergic and glutamatergic signaling as noted above, and recent studies have shown that acute alcohol consumption further disrupts this signaling (Zorumski et al., 2014), resulting in increased risk for other externalizing behavior including aggressive behavior while under the influence of alcohol (Miczek et al., 2015). Further, effects of chronic alcohol use on PFC functioning and exacerbation of EF deficits have been reported (Heinz et al., 2011). Together, these findings suggest EF impairments, and their associated neurobiological functions, may be an underlying risk mechanism for externalizing behaviors, but also highlight how externalizing behaviors can negatively impact these neurobiological systems and EF.

Genome-wide association study (GWAS) data have also been used to demonstrate the role of EF deficits in externalizing behavior using linkage disequilibrium score regression (LDSC; Bulik-Sullivan et al., 2015) and genomic structural equation modelling (Grotzinger et al., 2019), estimating genetic correlations between traits and evaluating the genetic basis of latent constructs hypothesized to explain the patterned covariances between studied traits, respectively. In particular, these studies have highlighted the genetic relations between EF task performance and externalizing behavior (Gustavson et al., 2023; Hatoum et al., 2023) and a latent addiction factor (Hatoum et al., 2022). Other research has explored the unique genetic correlations between externalizing behaviors in greater detail. For example, findings have found that alcohol use frequency is negatively correlated with ADHD and aggression, but ADHD is positively correlated with alcohol use quantity (Marees et al., 2020) and aggression (Demontis et al., 2023; Manchia & Fanos, 2017; Retz & Rösler, 2009). Combined, these studies indicate significant genetic overlap between externalizing behaviors and between EF and these behaviors at the genome-wide level. However, relatively little is known about their overlap at the level of genomic regions or biological pathways.

Thus, the current study employed genome-wide *and* local molecular genetic methods to refine our understanding of the biological mechanisms underscoring EF and externalizing phenotypes. First, the study examined whether the heritability of a trait or the co-heritability between two traits was enriched for genes expressed in particular tissues or stratified by other annotation types (e.g., conserved regions, DNase I-hypersensitive sites). Second, the study focused on smaller genomic segments to examine the genomic regions contributing to the covariation between traits and to identify plausible genes that may be influencing the associations between EF and externalizing traits. Given the reviewed evidence, it was hypothesized that these methods would consistently identify shared biological pathways between EF and alcohol use, aggression, and ADHD, particularly those related to dopaminergic or glutamate signaling and implicate genes preferentially expressed in the PFC and ACC tissues as activation of these brain regions has been associated with each phenotype. Additionally, we anticipated unique genetic overlap between EF and individual externalizing phenotypes.

## Methods

### GWAS

Analyses were conducted using GWAS summary statistics of aggression, alcohol consumption, AUD, ADHD, and EF. The ACTION consortium aggression GWAS summary statistics were derived from several cohort studies that totaled 87,485 children assessed for aggressive behavior via parent, teacher, or self-report (Ip et al., 2021). Alcohol consumption summary statistics were derived from a GWAS meta-analysis of the number of drinks consumed in a typical week (N=537,349 adults see Liu et al., 2019 for more details). AUD summary statistics were generated from a previously described GWAS meta-analysis (N=514,944 adults) using data from the Psychiatric Genetics Consortium (PGC; Walters et al., 2018), Million Veterans Program (Zhou et al., 2020), and FinnGen (FinnGen, 2021) cohorts (Spychala et al., 2024). AUD was measured as meeting either ICD-8, 9, or 10 (World Health Organization, 1967; 1988; 2004) criteria for AUD, or meeting DSM-IV (APA, 1994) criteria for alcohol dependence. ADHD summary statistics were derived from a meta-analysis of the iPSYCH (Bybjerg-Grauholm et al., 2020; Pedersen et al., 2018), PGC (Demontis et al., 2019), and deCODE cohorts (N=225,534 children; Demontis et al., 2023). ADHD diagnosis was defined as either meeting ICD-10 criteria for ADHD or receiving prescription medications to treat ADHD symptoms. Finally, EF summary statistics were generated from a GWAS of 427,037 adults from the UK Biobank sample (Hatoum et al., 2023) in which latent variable analysis was applied to tasks measuring different EF processes to extract a common EF factor. All GWAS summary statistics were conducted among individuals of European ancestry.

### Data Analysis

LDSC-SEG (Finucane et al., 2018) and Stratified-LDSC (S-LDSC; Finucane et al., 2015), were used to test whether the heritability of the traits and co-heritability between traits were enriched for genes expressed in particular tissues (e.g., fetal brain, ACC) or stratified by other annotation types (e.g., conserved regions, DNase I-hypersensitive sites) using the described summary statistics. For S-LDSC, a baseline model was estimated using 97 functional categories (e.g., Coding + 500bp, Promoter +500bp) from 24 publicly available annotations (e.g., coding, promoter). Additional models were estimated using 51 annotations developed from Roadmap chromatin data (i.e., DNase I-hypersensitive sites derived in the fetal brain; Roadmap Epigenomics Consortium et al., 2015). For LDSC-SEG, additional cell-type models using 205 expressed gene annotations from the Frank Lab (Pers et al., 2015) and 13 annotations from GTEx V8 (Aguet et al., 2020) were examined to identify enrichment in specific tissues (e.g., Angular gyrus, T cells). The Benjamini-Hochberg procedure (1995) was used to determine a false discovery rate (FDR) correction for each set of models to account for multiple testing across annotations. FDR-adjusted p-values were then calculated to evaluate statistical significance (P_FDR_<0.05).

As a complementary analysis, SUPERGNOVA (Zhang et al., 2021) was used to examine whether genetic covariation among the proposed traits could be attributed to shared enrichment of genes. SUPERGNOVA estimates local genetic covariation among pairs of phenotypes (i.e., aggression and ADHD) through weighted least squares regression for each independent LD block across the genome (2,353 total blocks). Significant results were then examined further by identifying potentially influential genes and genetic variants in those areas which have previously been associated with similar traits using the UCSC Genome Browser (Perez et al., 2025). Again, FDR-adjusted p-values (P_FDR_<0.05) were used for each pair of phenotypes to account for multiple testing across genomic regions.

Similarly, Local Analysis of [co]Variant Association tests (LAVA; Werme et al., 2022) was used to estimate local genetic covariation among pairs of phenotypes across 2,495 semi-independent genomic regions. First, LAVA was used to identify regions contributing to the overall heritability of each individual phenotype. A Bonferroni correction was employed to identify significant heritability of individual genomic regions for each trait (N=2,495 loci tested; p<2×10^-5^). Second, among pairs of traits demonstrating significant heritability at a genomic region, bivariate local genetic correlations were conducted. FDR-adjusted p-values used to evaluate statistical significance (P_FDR_<0.05).

Finally, the pairwise GWAS approach (GWAS-pw; Pickrell et al., 2016) was used to evaluate whether individual genomic segments (N=1,703) provided evidence of pleiotropy between pairs of traits. This approach simultaneously tests four models. Models 1 and 2 test the probability that a region contains a causal variant of the first or second phenotype, respectively. Model 3 examines the probability that the region contains a causal variant of both phenotypes. Model 4 examines the probability that the region contains two causal variants, each associated with a different phenotype. As the main aim was to identify pleiotropic regions, the focus was on the probabilities for Model 3. Pleiotropic regions were identified as those exhibiting a posterior probability ≥0.9 for Model 3, and thus, <0.1 for the remaining models.

## Results

### Stratified-LDSC

Across EF and externalizing phenotypes, S-LDSC identified enrichment in DNase hypersensitive sites assessed in the fetal brain and in genomic regions impacted by H3K4me1 epigenetic histone marks derived from the cortex, which are associated with gene expression (Bae & Lesch, 2020; P_FDR_< 0.05). Across alcohol consumption, AUD, ADHD, and EF, enrichment was identified in genomic regions conserved across species (P_FDR_<0.001). Enrichment in regions subject to negative selection was also identified for alcohol phenotypes and EF (P_FDR_<7.2×10^-6^) and approached significance for aggression and ADHD (P_FDR_>0.09).

Specific to EF and alcohol use phenotypes, alcohol consumption and AUD shared enrichment with EF in genomic regions impacted by H3K27ac epigenetic histone marks derived from the angular gyrus and DLPFC (P_FDR_<0.04), H3K4me3 epigenetic histone marks derived from the cingulate gyrus and DLPFC (P_FDR_<0.01), and H3K9ac epigenetic histone marks derived from the DLPFC (P_FDR_<0.01). Both H3K27ac and H3K9ac marks, when co-occurring with H3K4me3 marks, are associated with transcription initiation and increased gene activation and expression (Gates et al., 2017; Zhao et al., 2021). Alcohol consumption also shared enrichment with EF in genomic regions impacted by H3K9ac epigenetic histone marks derived from the anterior caudate (P_FDR_<0.001) and H3K27ac epigenetic histone marks derived from the cingulate gyrus (P_FDR_<0.001). ADHD and aggression did not demonstrate any unique sharing of enrichment with EF.

Outside of their relations with EF, alcohol consumption and ADHD shared enrichment in genomic regions characterized by H3K36me3 epigenetic histone marks (P_FDR_< 0.024), bivalent flanking transcription start sites (TSS: P_FDR_<0.05), and CpG content of promoters (P_FDR_<0.006).

### LDSC-SEG

For EF, LDSC-SEG cell-type analyses indicated significant enrichment after multiple testing corrections for genes expressed in 23 central nervous system tissues (P_FDR_<1.23x^10-8^). Cell-type analyses did not yield any significant findings for the other phenotypes after multiple testing corrections. However, analyses using the GTEx annotations indicated significant enrichment among genes expressed in the ACC (P_FDR_<0.02) and frontal cortex (P_FDR_<0.02) across all traits (**Figure 1**). Additionally, for EF, AUD, and alcohol consumption, there was significant enrichment for genes in 11 other brain tissues (e.g., amygdala, hippocampus) and ADHD demonstrated significant enrichment in 10 of those 11 additional tissues.

**Figure 1.**
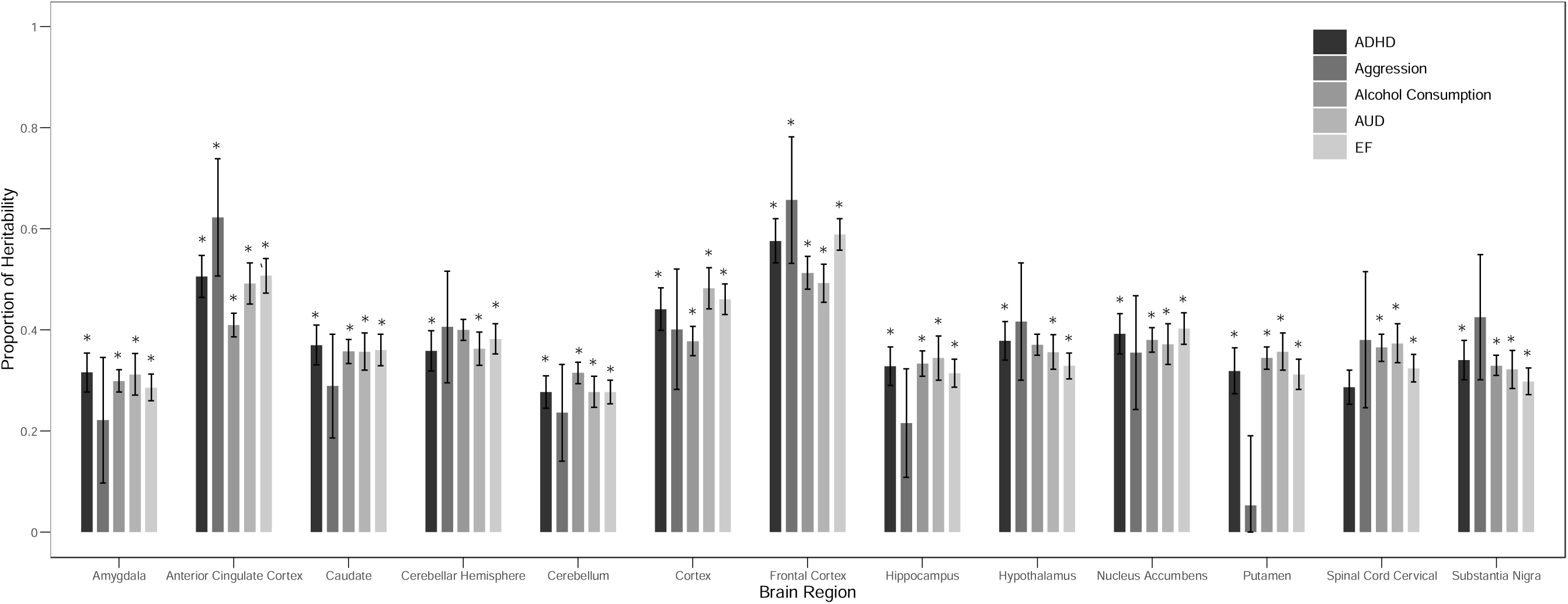
Note. Partitioned heritability estimates for ADHD, aggression, alcohol consumption, AUD, and EF from LDSC-SEG enrichment analyses using GTEx8 annotations. *indicates significant enrichment after FDR corrections. LDSC-SEG = Linkage-Disequilibrium Score Regression in Specifically Expressed Genes; ADHD = attention-deficit/hyperactivity disorder; AUD = alcohol use disorder; EF = executive function.

### SUPERGNOVA

EF showed 35 instances of significant covariation with externalizing phenotypes across 31 genomic regions (**Table 1**). Nineteen of these were with alcohol consumption, 12 were with ADHD, 3 were with AUD, and 1 was with aggression. Of note, there were several regions in which at least one phenotype showed localized genetic covariation with multiple other traits, each of which are described in detail below.

**Table 1.** Region Level Covariation Identified by SUPERGNOVA.

| Trait 1 | Trait 2 | Chromosome | Start | End | Rho | P<sub>FDR</sub> |
| --- | --- | --- | --- | --- | --- | --- |
| ADHD | EF | 1 | 43365565 | 44482312 | -0.0001 | **0.002** |
| ADHD | Aggression | 1 | 43365565 | 44482312 | 0.0005 | **0.01** |
| Aggression | EF | 1 | 205449511 | 206705609 | -0.0002 | **0.000002** |
| Alcohol Consumption | EF | 1 | 205449511 | 206705609 | -0.0001 | **0.004** |
| ADHD | EF | 1 | 242700803 | 244105053 | -0.0001 | 0.004 |
| AUD | EF | 2 | 73174848 | 74121000 | -0.00009 | **0.004** |
| Alcohol Consumption | EF | 2 | 73174848 | 74121000 | -0.0001 | **0.02** |
| ADHD | EF | 2 | 103264434 | 104481488 | -0.0002 | 0.003 |
| ADHD | Aggression | 2 | 114848115 | 116770829 | 0.0002 | 0.01 |
| ADHD | Alcohol Consumption | 2 | 163359084 | 165200324 | 0.00001 | 0.001 |
| ADHD | EF | 2 | 170973461 | 172086127 | -0.0001 | 0.03 |
| AUD | EF | 2 | 225833758 | 227531180 | 0.0001 | 0.03 |
| Alcohol Consumption | EF | 3 | 38080101 | 38723967 | -0.00005 | 0.02 |
| Alcohol Consumption | EF | 3 | 85093629 | 86734415 | -0.0001 | 0.02 |
| Alcohol Consumption | Aggression | 3 | 157312028 | 159272972 | 0.0002 | **0.003** |
| ADHD | Aggression | 3 | 157312028 | 159272972 | 0.0003 | **0.01** |
| ADHD | Alcohol Consumption | 4 | 103388441 | 104802530 | -0.0001 | 0.009 |
| Alcohol Consumption | EF | 5 | 11170703 | 13603734 | 0.0001 | 0.004 |
| ADHD | EF | 5 | 90504842 | 93602613 | 0.0002 | 0.003 |
| Alcohol Consumption | Aggression | 5 | 144584265 | 145803004 | 0.0001 | 0.04 |
| Alcohol Consumption | EF | 6 | 11235682 | 12183952 | -0.00009 | 0.004 |
| Alcohol Consumption | Aggression | 7 | 68562932 | 69806895 | 0.0001 | 0.0005 |
| ADHD | EF | 7 | 131857419 | 134306672 | -0.0003 | 0.001 |
| Alcohol Consumption | EF | 8 | 63711348 | 65193342 | -0.0001 | 0.03 |
| ADHD | EF | 8 | 66025862 | 67192642 | 0.0001 | 0.02 |
| Alcohol Consumption | EF | 8 | 92344138 | 94269630 | 0.0001 | 0.0001 |
| Alcohol Consumption | EF | 8 | 116095815 | 117130004 | 0.00004 | 0.03 |
| Alcohol Consumption | EF | 9 | 14714681 | 16054406 | -0.0001 | 0.004 |
| Alcohol Consumption | EF | 9 | 19456098 | 20324812 | -0.00009 | 0.02 |
| ADHD | Alcohol Consumption | 10 | 86181461 | 87571563 | 0.0001 | 0.006 |
| ADHD | EF | 11 | 16790799 | 18788126 | -0.0001 | 0.03 |
| ADHD | Alcohol Consumption | 11 | 27019873 | 28741185 | 0.0002 | **0.0009** |
| ADHD | EF | 11 | 27019873 | 28741185 | -0.0001 | **0.03** |
| Alcohol Consumption | EF | 11 | 27019873 | 28741185 | -0.00008 | **0.03** |
| Alcohol Consumption | EF | 11 | 113105405 | 113958177 | -0.0002 | **0.0001** |
| ADHD | Alcohol Consumption | 11 | 113105405 | 113958177 | 0.0002 | **0.00001** |
| Alcohol Consumption | EF | 12 | 2176265 | 2886299 | -0.0001 | 0.0001 |
| ADHD | Aggression | 14 | 33036081 | 34769821 | 0.0004 | 0.004 |
| ADHD | EF | 15 | 46294385 | 47000090 | -0.0001 | 0.004 |
| Alcohol Consumption | Aggression | 15 | 49078966 | 50004817 | 0.0001 | 0.001 |
| Alcohol Consumption | EF | 16 | 27446054 | 29023966 | 0.0001 | 0.000003 |
| Alcohol Consumption | EF | 16 | 29694822 | 31380596 | -0.0001 | 0.001 |
| ADHD | EF | 18 | 55090919 | 55808073 | -0.0001 | 0.03 |
| Alcohol Consumption | EF | 19 | 48697961 | 49485758 | 0.00008 | 0.03 |
| ADHD | Aggression | 20 | 19708351 | 21676556 | 0.0005 | 0.0000008 |
| ADHD | EF | 20 | 34892880 | 35892530 | -0.00007 | 0.03 |
| Alcohol Consumption | EF | 21 | 39250321 | 40115483 | -0.00006 | 0.03 |
| AUD | EF | 21 | 40116044 | 41095241 | 0.0001 | **0.001** |
| Alcohol Consumption | EF | 21 | 40116044 | 41095241 | 0.00007 | **0.03** |

*Note.* SUPERGNOVA indicated significant covariation between pairs of traits at the above regions. Regions with significant covariation between multiple pairs of traits is indicated in bold. Rho= estimation of local genetic covariation; AUD= alcohol use disorder, ADHD= attention-deficit/hyperactivity disorder; EF= executive functioning.

First, there was negative local genetic covariation between EF and ADHD, and positive local genetic covariation between ADHD and aggression at chromosome 1 (43365565: 44482312). This region contains the *HYI* and *SLC6A9* genes and variants that were previously associated with externalizing behavior (Demontis et al., 2021; Karlsson Linnér et al., 2021), general cognitive ability/intelligence (Davies et al., 2018; Savage et al., 2018), and educational attainment (Okbay, et al., 2022).

Second, negative genetic covariation was demonstrated between EF and alcohol consumption and EF and aggression at a second region of chromosome 1 (205459511: 206705609). This region contains the *NUCKS1* and *PM20D1* genes and variants that were previously associated with alcohol consumption (Saunders et al., 2022), cortical thickness (Shadrin et al., 2021), reaction time (Davies et al., 2018), and Parkinson’s disease (Kim et al., 2023).

Third and fourth, EF also demonstrated significant covariation with alcohol consumption and AUD on chromosome 2 (73174848: 74121000) and chromosome 21 (40116044: 41095241). The former region contains the *FBXO41* gene and both regions contain variants associated with cognitive ability (Davies et al., 2018; Homann et al., 2022), intelligence (Savage et al., 2018; Williams et al., 2023), smoking behavior (Liu et al., 2019; Saunders et al., 2022), and externalizing behavior (Karlsson Linnér et al., 2021).

Fourth and fifth, local genetic covariation was demonstrated between EF, ADHD, and alcohol consumption at an initial location on chromosome 11 (27019873:28741185) and between EF and alcohol consumption and between alcohol consumption and ADHD at a second location on chromosome 11 (113105405:113958177). This first region contains the *BDNF* and *METTL15* genes and variants that were previously associated with externalizing behavior (Karlsson Linnér et al., 2021 Saunders et al., 2022; Xu et al., 2023), along with general cognitive ability (Lee et al., 2018), and brain morphology (van der Meer et al., 2022). The latter region contains the *DRD2* gene, which is consistently associated with alcohol consumption (Deak & Johnson, 2021). It also contains the *NCAM1* gene and several variants previously associated with neuroticism (Kim et al., 2023), depression (Meng et al., 2024), and well-being (Baselmans et al., 2019).

Local genetic covariation was also demonstrated between the externalizing phenotypes independent of EF. ADHD demonstrated 10 significant instances of genetic covariation with the other externalizing phenotypes, alcohol consumption demonstrated nine instances, and aggression demonstrated eight instances. AUD did not demonstrate any significant covariation with the other externalizing phenotypes.

### LAVA

Across the five phenotypes and 2495 tested loci, 11,952 univariate heritabilities were estimated of which 6,115 survived multiple testing corrections (alcohol consumption=2,466, aggression=1906, AUD=930, EF=429, ADHD=384). Among phenotype pairs demonstrating significant heritability at a shared locus, 5,667 bivariate correlations were calculated. Of these, 272 survived FDR multiple testing corrections, and 22 loci demonstrated significant bivariate correlations with multiple pairs of phenotypes. Forty of these 272 significant correlations included EF. Of those, sixteen were correlations with alcohol consumption, and eight were correlations with AUD, ADHD, and aggression, respectively.

The remaining 232 significant correlations were between externalizing behaviors independent of EF. There were 116 correlations between alcohol consumption and AUD, 85 between alcohol consumption and aggression, and 13 between alcohol consumption and ADHD. Aggression demonstrated 12 significant correlations with AUD and ADHD, and there were 6 correlations between AUD and ADHD.

### GWAS-pw

Thirteen regions demonstrated significant evidence of pleiotropy in relation to EF and alcohol consumption (Model 3: PPA > 0.9) and eight regions demonstrated significant pleiotropy in relation to EF and AUD, including four of the same regions identified for EF and alcohol consumption (**Table 2**). Nine pleiotropic regions were also identified for EF and ADHD, one of which was a region identified to be pleiotropic for AUD and EF (chromosome 15: 90476610– 92164104). No significant pleiotropic regions were identified between EF and aggression.

**Table 2.** Pleiotropic Regions Between Traits Identified by GWAS-PW.

| Trait 1 | Trait 2 | Chromosome | Start | End | PPA |
| --- | --- | --- | --- | --- | --- |
| ADHD | EF | 1 | 5914245 | 7245669 | 0.98 |
| AUD | Aggression | 1 | 71684831 | 74326378 | 0.91 |
| Alcohol Consumption | EF | 1 | 204682011 | 205922403 | 0.95 |
| ADHD | EF | 2 | 10298963 | 12418752 | 0.94 |
| **Alcohol Consumption** | **EF** | **2** | **142518602** | **144517069** | **0.98** |
| **AUD** | **EF** | **2** | **142518602** | **144517069** | **0.99** |
| Alcohol Consumption | EF | 3 | 139954597 | 141338641 | 0.97 |
| **Alcohol Consumption** | **EF** | **4** | **100679579** | **103220206** | **0.99** |
| **AUD** | **EF** | **4** | **100679579** | **103220206** | **0.99** |
| **Alcohol Consumption** | **EF** | **4** | **103224214** | **105303155** | **0.96** |
| **AUD** | **EF** | **4** | **103224214** | **105303155** | **0.94** |
| Alcohol Consumption | ADHD | 5 | 87391024 | 88890628 | 0.98 |
| Alcohol Consumption | EF | 6 | 97842747 | 100629461 | 0.97 |
| AUD | EF | 7 | 4575430 | 5415190 | 0.93 |
| AUD | EF | 7 | 113712387 | 116779594 | 0.91 |
| ADHD | EF | 8 | 11280885 | 13491197 | 0.97 |
| Alcohol Consumption | EF | 8 | 20060856 | 21622342 | 0.96 |
| AUD | ADHD | 8 | 76456641 | 79132323 | 0.91 |
| AUD | EF | 8 | 143045846 | 144236013 | 0.96 |
| Alcohol Consumption | EF | 9 | 18661683 | 20455527 | 0.97 |
| Alcohol Consumption | EF | 10 | 100668400 | 102948934 | 0.98 |
| ADHD | EF | 11 | 28481593 | 29969601 | 0.91 |
| AUD | ADHD | 11 | 55083434 | 58455947 | 0.91 |
| **Alcohol Consumption** | **EF** | **12** | **51776494** | **53038588** | **0.93** |
| **AUD** | **EF** | **12** | **51776494** | **53038588** | **0.96** |
| ADHD | EF | 14 | 28467299 | 29969601 | 0.92 |
| Alcohol Consumption | EF | 14 | 71136946 | 72888427 | 0.95 |
| Alcohol Consumption | EF | 15 | 80861289 | 84259565 | 0.94 |
| **ADHD** | **EF** | **15** | **90475963** | **92164104** | **0.94** |
| **AUD** | **EF** | **15** | **90475963** | **92164104** | **0.91** |
| ADHD | EF | 16 | 52036960 | 53375951 | 0.95 |
| Alcohol Consumption | EF | 18 | 913491 | 1910854 | 0.97 |
| ADHD | EF | 20 | 25344231 | 29969601 | 0.91 |
| ADHD | EF | 22 | 22359861 | 23712520 | 0.95 |

*Note.* GWAS-PW identified significant pleiotropy between pairs of traits at the above regions. Regions with significant pleiotropy between multiple pairs of traits is indicated in bold. PPA= posterior probability that the region contains a variant influencing both traits; AUD= alcohol use disorder, ADHD= attention-deficit/hyperactivity disorder; EF= executive functioning.

Across the three methods examining genomic overlap at the regional level (i.e., SUPERGNOVA, LAVA, and GWAS-pw), 23 overlapping regions located on 12 different chromosomes were identified by two or more of the methods, with two regions identified in all methods (**Figure 2**).

**Figure 2.**
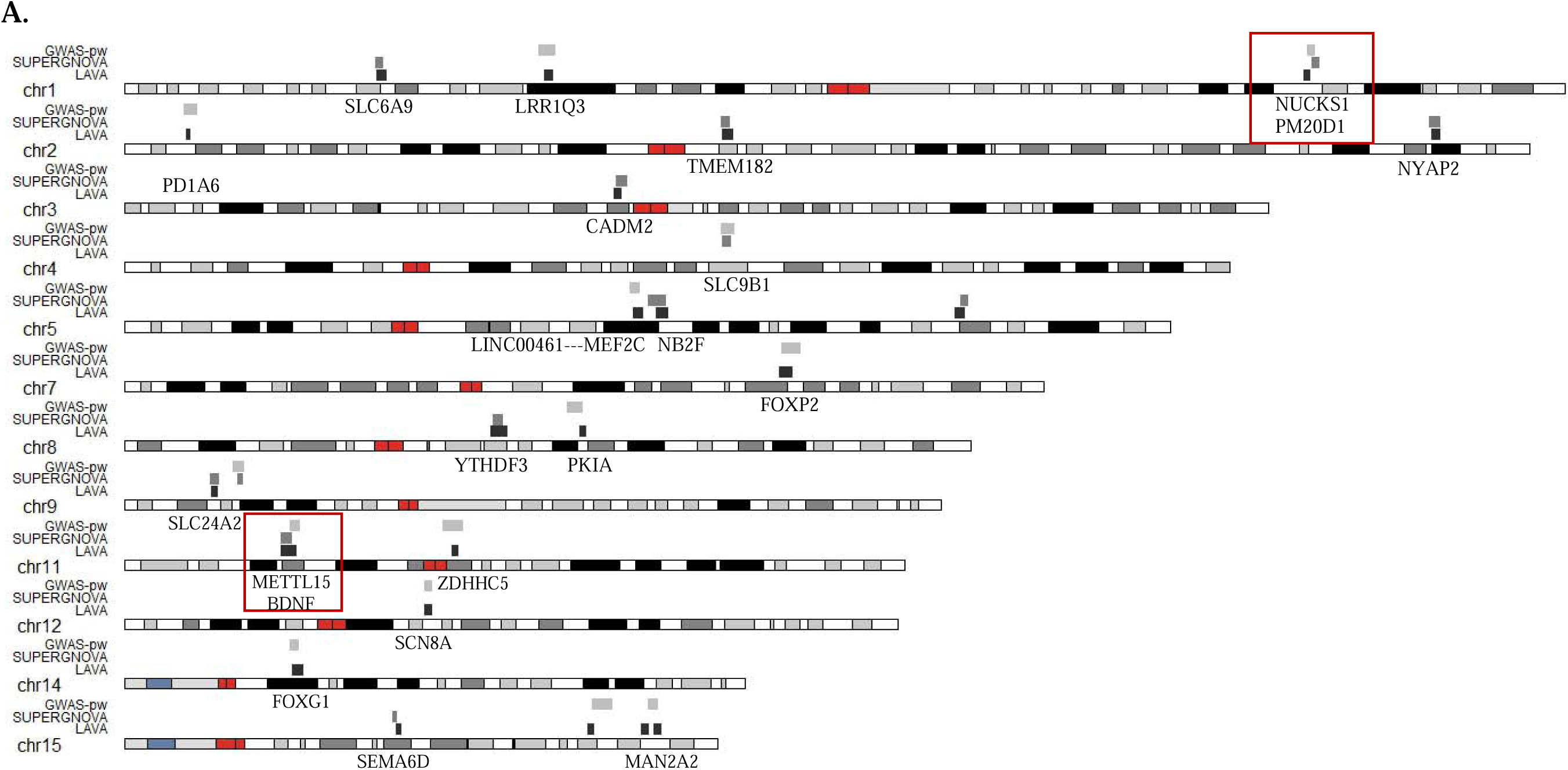
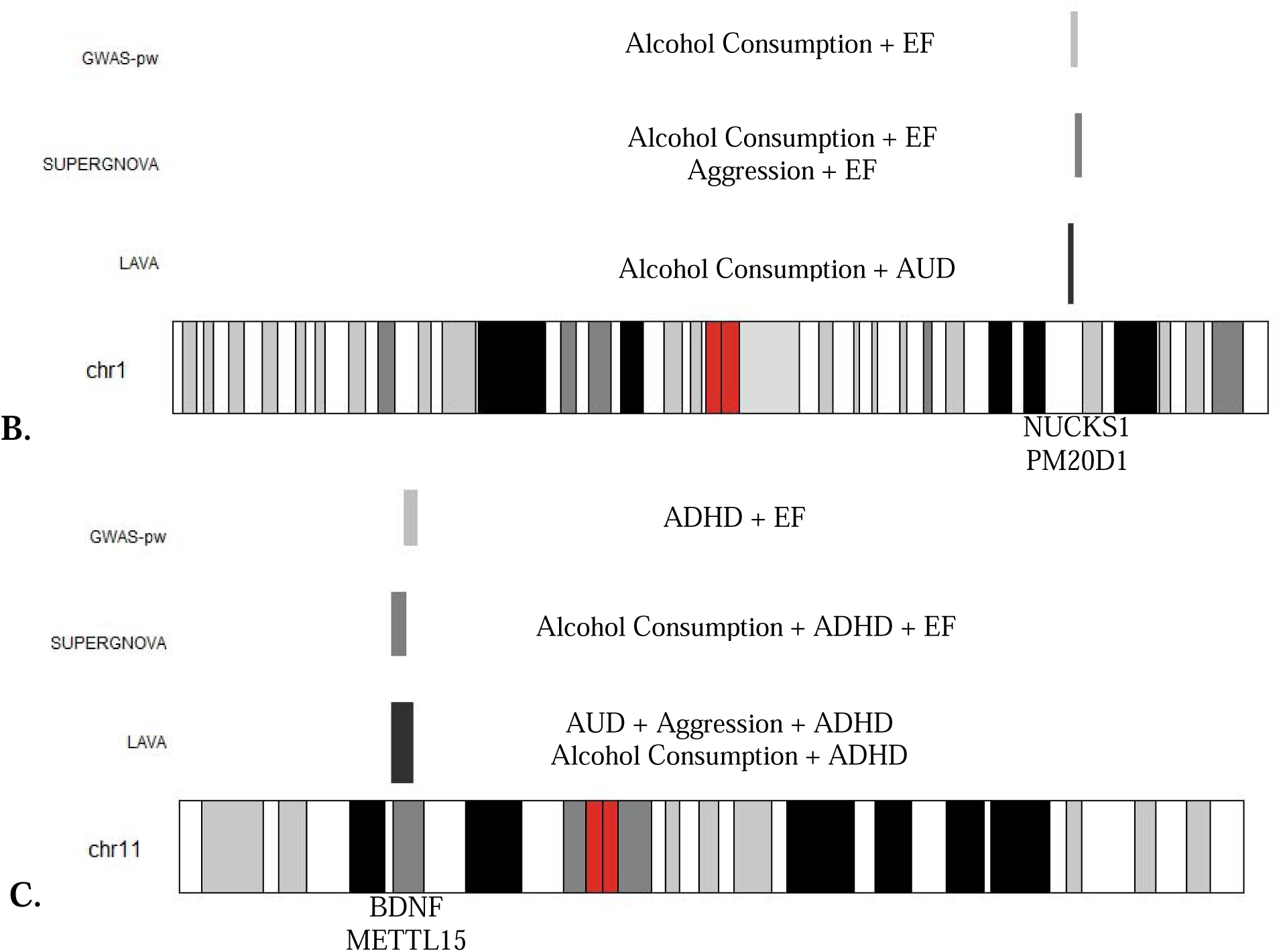
Note. Karyotype plots of regions evidencing correlations between multiple phenotypes across the three regional methods (e.g., SUPERGNOVA, LAVA, GWAS-pw). Regions encompassed by a red box were identified in all three methods. Mapped genes to these regions are labeled below the region in black. A. Regions identified in at least two methods. Regions encompassed by a box indicate they were identified in all regional methods. B. Zoomed in karyotype plot of the region on chromosome 1 identified in all three methods. Phenotypes significantly correlated after multiple testing corrections at the indicated region are labeled next to the associated method. C. Zoomed in karyotype plot of the region on chromosome 11 identified in all three methods. Phenotypes significantly correlated after multiple testing corrections at the indicated region are labeled next to the associated method. chr = chromosome; LAVA = Local Analysis of [co]Variant Association; GWAS-pw = pairwise GWAS; ADHD = attention-deficit/hyperactivity disorder; AUD = alcohol use disorder; EF = executive function.

## Discussion

The current study employed several molecular genetics methods to explore the hypotheses that EF deficits represent a shared underlying mechanism for the externalizing phenotypes of alcohol use, AUD, ADHD, and aggression but also that EF may exhibit unique relations with these phenotypes. Findings related to each of these hypotheses from both genome-wide enrichment and regional methods are highlighted.

### Executive Functioning as a Shared Risk Factor for Externalizing Behaviors

Among methods that parsed the genetic influences underlying EF and externalizing phenotypes according to genes expressed in particular tissues or stratification along other annotation types, there was evidence for enrichment of genetic signals in DNase hypersensitive sites and H3K4me1 regions, both of which are associated with gene expression (Chen & Chen, 2019 and Bae & Lesch, 2020, respectively). There was also evidence for significant enrichment among genes expressed in the ACC and frontal cortex. In addition to enrichment analyses, approaches for identifying individual genes or genomic segments that contribute to the shared etiology among EF and externalizing phenotypes were employed. Combining results across all regional methods regions on chromosomes 1, 11, and 15 evidenced support for an association with EF and all externalizing behaviors. Each of these regions contains genes previously associated with cognitive functioning and/or psychopathology that could explain the observed relations.

The region on chromosome 1 (43365565-45167235) contains *SLC6A9,* which encodes for a glycine transporter that influences the interaction of glutamate and dopamine in the brain (de Bartolomeis et al., 2020; Rodek et al., 2022). The region on chromosome 11 contains *BDNF* which encodes for brain derived neurotrophic factor, a protein involved in promoting the survival of cells, synaptic transmission in the brain, and memory (Miranda et al, 2019), and *METTL15*, which encodes for a protein associated with methylation of mitochondrial ribosomal RNA (Safran et al., 2021). Finally, the region on chromosome 15 (89385188-92675405) contains *SLC03A1*, which encodes for a transporter protein enriched in cortical grey matter (Safran et al., 2021). These genes have been previously associated with externalizing behavior (Demontis et al., 2021; Karlsson Linnér et al., 2021; Saunders et al., 2022) and cognitive-related phenotypes (Davies et al., 2018, Okbay et al., 2022). The chromosome 1 findings are also consistent with our hypotheses that glutamatergic and dopaminergic transmission represent shared biological mechanisms underpinning EF and externalizing behavior and is concordant with prior human and animal studies demonstrating the relevance of these processes in the neurobiology of these phenotypes (Aultman & Moghaddam, 2001; Davidson et al., 2000; Huang et al., 2019; Yan & Rein, 2022). While this represents just a snapshot of the relevant genes within these regions, their relevance to cognition provides evidence supporting their relevance to EF and EF’s role as a shared risk mechanism underlying the externalizing spectrum.

Though these findings provide biological support for the hypothesis that EF deficits serve as risk factors for externalizing behaviors, there is a question of whether this risk extends beyond the externalizing spectrum. For example, the significant expression patterns described above have been observed for other types of psychopathology (Howard et al., 2019), and some of the described genomic regions have shown associations with other psychiatric disorders (Baselmans et al., 2019; Curtis, 2022). Similar findings also been reported in the phenotypic literature. For example, Shanmugan et al. (2016) showed that different types of psychopathology (i.e., anxiety, psychosis, fear, and externalizing behavior) demonstrate similar deficits in activation of executive regions, suggesting that EF may be a transdiagnostic risk factor. Consistent with such evidence, bifactor models have demonstrated strong associations between EF and a general P factor and minimal associations with an externalizing-specific factor (Bloemen et al., 2018), a finding that has been replicated in behavior genetic (Harden et al., 2020) and structural neuroimaging studies (Romer & Pizzagalli, 2022). The identification of genes and/or enrichment associated with both externalizing and other forms of psychopathology in the current study suggests EF may be related to externalizing phenotypes through general psychopathology mechanisms.

### Executive Functioning as a Risk Factor for Individual Externalizing Behaviors

The current study also sought to identify unique genetic relations between EF and each of the externalizing behaviors. Across methods, alcohol consumption demonstrated the strongest relations with EF. For example, EF showed the greatest shared enrichment with alcohol consumption in genomic regions impacted by epigenetic histone marks. Additionally, alcohol consumption demonstrated associations with EF across genomic regions not identified in relations with other externalizing behaviors. Notably, two of these regions were identified across multiple methods, chromosome 8 (20060856 - 21650453) and chromosome 9 (18661683 - 20455527). Each of these regions contains genes previously identified in GWAS of cognitive-related factors, risk tolerance, and substance use (e.g., Debette et al., 2015; Gelernter et al., 2015; Karlsson Linnér et al., 2019). The former region contains *GFRA2*, responsible for encoding the protein associated with glial cell-derived neurotrophic factor (Safran et al., 2022), involved in the protection of dopaminergic neurons from damage (Airaksinen & Saarma, 2002), and has previously been implicated as a target for reducing alcohol consumption in rat models (He et al., 2005). The latter region contains *SLC24A2,* which has been associated with deficits in long term memory in mice (Li et al., 2006) and alcohol consumption in humans (Leger et al., 2024). Together, these findings suggest biological pathways specific to the relations between EF and alcohol consumption, indicating unique relations between EF and individual externalizing behaviors may be present.

These findings highlight a second potential implication of the present study. Namely, while EF may be broadly related to externalizing behavior and/or psychopathology, individual loci may also be uniquely associated with individual externalizing behaviors potentially through distinct relations with individual EF component processes and distinct neurobiological systems. It is also possible that some components of EF are more or less associated with individual externalizing behaviors (Weyandt et al., 2014), such as alcohol consumption, and thus would show different patterns of association with an EF latent factor. This is consistent with the previously discussed evidence suggesting differential patterns of brain activation during response inhibition and delay discounting tasks between ADHD and conduct disorder relative to substance misuse (Castellanos-Ryan et al., 2014). However, the current study did not explore associations with specific EF processes, and thus, cannot differentiate between these possibilities.

### Relations Between Externalizing Behaviors Independent of Executive Function

In addition to the unique relations between externalizing phenotypes and EF, there was evidence for relations between externalizing phenotypes independent of EF. For example, alcohol consumption and ADHD shared unique patterns of enrichment in genomic regions impacted by H3K36me3 epigenetic histone marks, and the CPG content of promoters. Similar results were obtained from the regional methods, including shared relations between alcohol consumption, aggression, and ADHD to a region of chromosome 5 (143605909–145803004). Thus, a final implication derived from the current study is that while deficits in EF may be an underlying mechanism of externalizing behavior, shared genetic and biological underpinnings with EF do not fully explain the relations between these behaviors. For example, impulsivity and EF, while not highly correlated with one another, are independently associated with externalizing behavior (Friedman et al., 2020), suggesting EF explains only certain facets of externalizing behavior. Thus, future studies applying the present study methods to other hypothesized risk mechanisms underlying externalizing behaviors are warranted.

## Limitations and Conclusion

Though important, the described implications should be interpreted considering several limitations of the current study. First, the sample size of the aggression GWAS (N=87,485) was considerably smaller than the other GWAS used in the present study and resulted in no identifiable genome-wide significant SNPs for this phenotype (Ip et al., 2021). This ultimately reduced the power to detect enrichment and associations with other phenotypes at the regional level (Werme et al., 2022). Additionally, the genomic regions defined by the local methods were quite large and differed in their boundaries across methods, making it difficult to pinpoint the exact variants and genes and associated biological mechanisms contributing to the relations between EF and externalizing phenotypes. Finally, the exclusive use of GWAS summary statistics derived from individuals of European ancestry represents a significant limitation of the current study. This was unavoidable, however, as the methods utilized require all summary statistics be derived from the same ancestry group, and the EF and aggression GWASs were exclusively conducted in European ancestry samples (Finucane et al., 2018; Werme et al., 2022; Zhang et al., 2021). Future work should conduct multi-ancestry GWAS to explore whether these findings generalize across ancestry groups.

Despite these limitations, the reported findings provide genetic evidence supporting EF as an underlying mechanism of externalizing behavior while suggesting that individual externalizing behaviors may also have unique relations with EF. Future research should focus on disentangling EF as a mechanism of externalizing behaviors and psychopathology more broadly by evaluating shared biological pathways among EF and other forms of psychiatric disorders (e.g., depression, schizophrenia). Additionally, as EF is a complex phenotype derived of multiple component processes (Diamond, 2013; Friedman & Miyake, 2017; Miyake et al., 2000), similar analyses could also explore the genetic relations between EF component processes and individual externalizing behaviors, as well as other forms of psychopathology, to facilitate the identification of transdiagnostic targets for future intervention and prevention efforts.

## Data Availability

The alcohol consumption GWAS summary statistics were obtained from https://conservancy.umn.edu/items/ca7ed549-636b-41c0-ae79-97c57e266417. PGC alcohol dependence GWAS summary statistics were obtained from the PGC website (https://www.med.unc.edu/pgc/). Million Veteran Program GWAS summary statistics were obtained through the Database for Genotypes and Phenotypes (dbGaP; Study Accession: phs001672). FinnGenR5 ICD-based AUD GWAS data were obtained from https://www.finngen.fi/en/access_results. The executive functioning GWAS summary statistics are available at http://ftp.ebi.ac.uk/pub/databases/gwas/summary_statistics/GCST90162001-GCST90163000/GCST90162547. The aggression GWAS summary statistics are available at https://tweelingenregister.vu.nl/eagle-gwa-meta-analyses-summary-results. The ADHD GWAS summary statistics were obtained from https://pgc.unc.edu/for-researchers/download-results/

## Acknowledgments

This study made use of summary statistics data from a number of sources. First, this research used summary data from the UK Biobank, a population-based sample of participants whose contributions we gratefully acknowledge. Second, this study used GWAS summary statistics data from the fifth release of the FinnGen study. We thank the participants and investigators of the FinnGen study. Third, this research also used summary data from the Psychiatric Genomics Consortium Substance Use Disorders (PGC-SUD) working group. PGC–SUD is supported by funds from NIDA and NIMH to MH109532 and, previously, had analyst support from NIAAA to U01AA008401 (COGA). PGC–SUD gratefully acknowledges its contributing studies and the participants in those studies, without whom this effort would not be possible. Fourth, this research used data from the Million Veteran Program and was supported by funding from the Department of Veterans Affairs Office of Research and Development, Million Veteran Program Grant nos. I01BX003341 and I01CX001849; and the VA Cooperative Studies Program study, no. 575B. This publication does not represent the views of the Department of Veterans Affairs or the United States Government. Fifth, this study used summary statistics data from the ACTION consortium. We gratefully acknowledge the ACTION participants for their contributions. Sixth, this study used summary statistics data from cohorts contributing to GSCAN Phase 1. We gratefully acknowledge the participants of these cohorts for their contribution. Finally, this research used summary statistics data from the iPSYCH consortium, deCODE, the ADHD working group of PGC (PGC-ADHD), Yale-Penn, Partners Biobank, and BioVU. We thank the participants of the iPSYCH, deCODE, PGC-ADHD, Yale-Penn, Partners Biobank, and BioVU cohorts for their contributions.

## Funding Statement

This work was supported by the National Institute on Alcohol Abuse and Alcoholism (KS, grant number: F31AA029948; AP, grant number: K01AA031724) and the National Institute on Drug Abuse (KS, PI: Arpana Agrawal and Jose Moron-Concepcion, grant number: T32DA007261; AP, PIs: Kathleen K. Bucholz and Jeremy T. Goldbach grant number: T32DA015035).

## Ethical Standards

The authors assert that all procedures contributing to this work comply with the ethical standards of the relevant national and institutional committees on human experimentation and with the Helsinki Declaration of 1975, as revised in 2008.

## Notes

### Competing Interest Statement

The authors have declared no competing interest.

### Author Declarations

All secondary data analysis of GWAS summary statistics and reference panels were considered exempt by the Institutional Review Board at the University of Missouri.

